# Performance of a Multisensor Smart Ring to Evaluate Sleep: In-Lab and Home-Based Evaluation Relative to Polysomnography and Actigraphy: Importance of Generalized Versus Personalized Scoring

**DOI:** 10.1101/2021.12.22.21268267

**Authors:** Michael A. Grandner, Zohar Bromberg, Aaron Hadley, Zoe Morrell, Arnulf Graf, Stephen Hutchison, Dustin Freckleton

## Abstract

**Study Objectives:** Wearable sleep technology has rapidly expanded across the consumer market due to advances in technology and increased interest in personalized sleep assessment to improve health and mental performance. In this study, we tested the performance of a novel device, the Happy Ring, alongside other commercial wearables, against in-lab polysomnography (PSG) and an at-home EEG-derived sleep monitoring device, the Dreem 2 Headband.

**Methods:** 36 healthy adults with no diagnosed sleep disorders and no recent use of medications or substances known to affect sleep pattern were assessed across 77 nights while wearing the Happy Ring, as well as a set of other consumer wearable devices. Subjects participated in a single night of in-lab PSG and 2 nights of at-home data collection. The Happy Ring includes sensors for skin conductance, movement, heart rate, and skin temperature. The Happy Ring utilized two machine-learning derived scoring algorithms: a “generalized” algorithm that applied broadly to all users, and a “personalized” algorithm that adapted to individual subjects’ data. Epoch-by-epoch analyses compared the wearable devices to both in-lab PSG and to the Dreem 2 EEG Headband (“Dreem 2 Headband”) at home.

**Results:** Compared to in-lab PSG, the “generalized” and “personalized” algorithms demonstrated good sensitivity (94% and 93%, respectively) and specificity (70% and 83%, respectively). Accuracy was 91% for “generalized” and 92% for “personalized” algorithms. The generalized algorithm demonstrated an accuracy of 67%, 85%, and 85% for light, deep, and REM sleep, respectively. The personalized algorithm was 81%, 95%, and 92% accurate for light, deep, and REM sleep, respectively.

**Conclusions:** The Happy Ring performed well at home and in the lab, especially regarding sleep detection. The personalized algorithm demonstrated improved detection accuracy over the generalized approach and other devices, suggesting that adaptable, dynamic algorithms can enhance sleep detection accuracy.

## INTRODUCTION

Personal sleep health is increasingly recognized as an important factor in physical and mental health, affecting one’s performance, response to stressors, and emotional regulation throughout the day^1–6^. Wearable technology provides opportunities to assess sleep health in real-world situations so that associations with relevant health and performance outcomes can be examined in the context of reliable, objective recordings in naturalistic settings^7–11^. This has proven useful, as wearable technology to assess sleep has been used to explore associations with cardiovascular disease^12,13^, metabolic dysregulation^12,14^, inflammation^15,16^, mental health^17,18^, physical performance^19^, and other outcomes, including mortality^20^.

As the technology develops, there is an increasing proliferation of wearable devices aimed at the consumer market^21,22^. Initially, these devices offered a high degree of scalability, but presented significant limitations on accuracy^23–25^. However, this gap has been closing recently, as studies increasingly demonstrate that newer consumer sleep-tracking devices can reliably estimate sleep-wake states with at least as much accuracy as standard scientific devices^26–29^. Improved sleep-wake prediction performance in consumer devices is likely related to the increased capabilities of physiologic signals in wearable technology^27,30^. Additionally, scoring algorithms have advanced, and now frequently include complex machine learning algorithms that may also improve sleep detection^31–33^.

For these reasons, standards for the validation^9,34^ and implementation^10,35^ of new technologies have been put forward for sleep predictions. Further, the concept of evaluating the performance of devices for sleep prediction has emerged as a strategy for addressing the limitations of more traditional validation approaches^10,36^. With the establishment of these standards, applications of novel sensors and scoring strategies have been called for^11^.

Accordingly, the present study evaluates the performance of a novel device both in the laboratory and at home, relative to a gold-standard comparator (in-lab polysomnography; PSG), an at-home EEG device (Dreem 2 Headband), and several other commercially available wearable devices. Specifically, this study aims to (1) quantify the accuracy of sleep detection relative to in-lab and at-home sleep assessment by evaluating sensitivity and specificity, (2) quantify the accuracy of sleep stage classification compared to in-lab and at-home sleep assessment by evaluating sensitivity and specificity, (3) quantify the relative performance of the investigational device compared to existing devices that have previously demonstrated good performance, and (4) evaluate the relative performance of two different machine learning approaches for sleep scoring.

## METHODS

### Participants

Participants were recruited through multiple strategies, including emailing past research study participants, and using word of mouth. Participants were eligible to participate if they: fell within the age range of 20 to 65 years of age, had no known or diagnosed sleep, mental health or significant medical disorders, were not pregnant, had no regular nicotine use within the past month, no habitual use of antidepressants, beta-blockers, stimulant medications, prescription pain medications, or anti-seizure medications within the past three months, had not used THC products within the past two weeks, and had a habitual sleep window and average nightly sleep within a normal range. A normal sleep window was defined as average bedtime after 9 pm and before 2 am and average wake-up time after 5 am and before 10 am. The study protocol was approved by the Solutions Institutional Review Board.

### Measures

The study device (“Happy Ring”) consisted of a finger-worn device that includes sensors for: (1) electrodermal activity (EDA), (2) 3-axis accelerometry, (3) photoplethysmography (PPG) -derived heart rate, and (4) skin temperature. EDA was measured using a medical grade 3-electrode impedance sensor, with data collected at 8 Hz. Raw data collected from the Happy Ring were streamed to an associated digital application through a data download process conducted each day of the study period, and sleep outcomes for each night of sleep were calculated offline using Python.

Accelerometry was measured using a smartphone-class microelectromechanical, multiaxial accelerometer embedded within the device, with data collected at 104 Hz. Data collected at 100 Hz from the PPG sensor provided the opportunity to assess heart rate, computed using a proprietary algorithm. Skin temperature was collected from the Happy Ring using a medical grade +/- 0.1 °C accuracy temperature sensor, data were collected at 1 Hz. Lastly, the internal clock of the unit computed time elapsed since the start and end of each sleep period. From these signals, data were merged into 30-sec epochs across all channels. Key metrics obtained from the device included epoch-by-epoch sleep and wake estimations, as well as time in bed (TIB), sleep latency (SL), wake after sleep onset (WASO), and total sleep time (TST; computed as TIB – SL – WASO). In addition, a proprietary algorithm was used to detect sleep stages, further classifying sleep as either “Deep” (Non-rapid eye movement (NREM); stage N3), “Light” (NREM stage N1 or N2), or REM. Sleep stages were determined based on a machine learning algorithm.

In-lab polysomnography (PSG) was accomplished using a Respironics Alice 6 diagnostic sleep system, with Natus Neurology Gold Cups as leads. The system used electroencephalogram (EEG; F3-A2,F4-A1,C3-A2,C4-A1,O1-A2,O2-A1), electromyogram amplifier module (EMG; chin) and electrooculogram (EOG; LOC-A2, ROC-A2) channels. In addition to 3 electrocardiogram (EKG) patches, chest and abdomen belts, and a thermistor and a nasal cannula. Scoring was performed by a certified polysomnographic technician, following the standard American Academy of Sleep Medicine scoring rules^37^.

At-home sleep assessment was performed using the Dreem 2 Headband to record electroencephalographic and other signals. The Dreem 2 Headband includes 5 dry-EEG sensors (O1, O2, FpZ, F7, F8), in addition to a 3D accelerometer to monitor movement and head position^38^. Participants wore the Dreem 2 Headband on two consecutive nights at home. Data were scored using the standard Dreem settings. Sleep stage predictions using the Dreem 2 Headband are provided automatically, with demonstrated accuracy in the range of individual scorers using PSG data^38^. Although the Dreem 2 Headband does not completely replace standard in-laboratory PSG, it does approximate laboratory PSG better than sleep wearables that use only peripheral signals^28,39^.

In addition to the Happy Ring, participants were also asked to wear up to four other sleep-tracking devices. First, participants were asked to wear the Actiwatch 2 from Philips Respironics. The Actiwatch is a gold-standard actigraphy device, with well-characterized validation data^7^. The Actiwatch uses a 3-axis accelerometer and scores sleep based on movement data. Data were extracted from the wristband at the conclusion of each trial using the computer-based Actiware software, and were scored using the standard algorithm, without additional hand-scoring. Second, participants were asked to wear a Fitbit device (Fitbit Charge 2, Fitbit Inc.). This commercially available device uses both accelerometry and PPG heart rate data to estimate sleep and wake^30^. Previous studies have shown that the Fitbit device is a valid and reliable tool for assessment of sleep, with strong agreement compared with PSG (sleep-wake sensitivity ≥0.93)^28,30,40,41^. Third, participants were asked to wear a Whoop device (Whoop 3.0, Whoop Inc.). This device is also commercially available and uses both movement and PPG data to estimate sleep^42,43^. Some previous studies have demonstrated that this device is accurate when compared to PSG (sleep-wake sensitivity ≥0.90)^42^. Finally, participants were asked to wear an Oura Ring (model V2). Like the Happy Ring, the Oura Ring is worn on the finger, and includes sensors for accelerometry and PPG heart rate data for the estimation of sleep^44^. Due to inventory constraints and a desire to maintain participant compliance to the study protocol, not all participants wore all devices during the three nights of data collection.

Devices were rotated between users to make sure the same number of nights of data were available for all devices. Data were collected from Fitbit Charge 2, Whoop Wristband, and Oura Ring devices through device-associated digital applications downloaded to an iPhone SE provided to the participant. All systems output hypnograms in 30 second windows, except for Oura, which outputs in 5 minute windows. Sleep windows for each device were auto-detected, with participants being instructed to put on the devices at least one hour before their anticipated bedtime.

Device placements for finger-based wearables were determined by best fit. Several device sizes were provided at the sleep lab to select the optimal size, and best fit fingers were selected for both the Happy Ring and the Oura Ring, prioritizing placement of both rings on the non-dominant hand. Additionally, the Actiwatch was placed on the right wrist, and the Whoop and Fitbit devices were both placed on the left wrist, with the Whoop placed proximal, and the Fitbit placed distal from the hand.

### Procedure

All participants participated in a three-night study which included one night in a sleep laboratory and two subsequent nights at home. Data were collected from participants over the course of two months between March 2021 and May 2021. In the lab, the sleep period was defined as lights off and lights on according to the PSG labels for the lab trials. Lights off were enforced by the lab technician based on a 60-minute window of each participant’s self-reported normal weekday bedtime. Lights on were marked based on each participants’ natural wake up, but were enforced if the wake up had not occurred by 7:00 AM local time. At home, participants were instructed to adhere to their regular sleep schedule, and the sleep period was defined as the start and end of the Dreem 2 Headband recordings. The start of each Dreem 2 headband recording was user-initiated within the associated digital application, and recording end was initiated by removing the device.

Preprocessing of Happy Ring signals included linear interpolation of missing data when the gap was less than 3 seconds. When the gap was larger, null /missing values were used. Signals were interpolated to 52 Hz, 8 Hz, 1 Hz, and 1 Hz for acceleration, EDA, heart rate, and temperature, respectively. The sleep lab and at-home sleep datasets were analyzed separately.

All data from the Happy Ring were analyzed in 30-second epochs. From these data, 211 features were extracted: 55 features from acceleration, 26 features from EDA, 103 features from heart rate, 26 features from temperature, and 1 feature from time elapsed. Examples of features are mean, standard deviation, range, and frequency of values and variables. To standardize values, they were converted to z-scores for each data sample, defined as a night of sleep.

### Determining Sleep-Wake and Sleep Stage Data with the Happy Ring

Personalized Happy algorithms were developed by being trained only on Happy Ring data to tailor output to the individual. A central part of the analysis was how the train-test split was defined., In other words, determining what time snippets of the data were used to learn the parameters of the classifier, and what snippets were used to infer sleep stages^45,46^. Two sets of metrics were used to quantify sleep stage prediction accuracy:

1. “Happy Generalized” was developed by doing a leave-one-out cross-validation^45,46^ at the level of the nights. This approach involved training on all but one subject’s data and then testing on that user, repeating across users. This approach tests the generalization ability of the sleep stage classifier across subject nights, with learning and inference done on different subject nights.
2. “Happy Personalized” was developed by training and testing on all subjects for different non-overlapping time windows, allowing for learning and inference on identical groups of subjects. This included a cross-validation where 80% of the data were used for training, and 20% of the data were reserved for testing. For testing, the segments for each night were stitched back together. This approach evaluates the personalization of the sleep stage classification.

If the data contained many subjects that had a wide range of physiological responses, both approaches would yield similar results. However, if the data were based on a small subset of subjects or nights, both approaches can give different results reflecting the data sampling bias, and not the inference quality^**45**,**46**^. Therefore, the Happy Generalized approach could underestimate the sleep stage prediction accuracy because training and testing data may be very different, whereas the Happy Personalized scoring method would give a more accurate representation of the prediction accuracy because the classifier learns from each subject night.

Both approaches inform on different aspects of sleep stage prediction. Happy Generalized can be interpreted as a generalized approach that does not take into consideration the evaluated subject. On the other hand, Happy Personalized used the evaluated subject for learning, providing a more individualized sleep prediction that adapts and learns from the user over time. In the context of our research, Happy Generalized validated the models by setting a baseline, and Happy Personalized improved upon the baseline by personalizing the sleep stage prediction process.

The training data exhibited a strong class imbalance (22% deep, 41% light, 27% REM and 10% awake samples across all nights). To overcome this, the training data were randomly under-sampled. A random forest classifier was trained and determined the optimal number of estimators as 10 using a cross-validated grid search. The classifier provided a probability-of-class membership for each sample in the unbalanced testing set. Then, post-processing of interpolation and Gaussian smoothing was applied to this probability distribution before making a categorical prediction by finding the location of the maximum of this probability distribution. Finally, *ad hoc* heuristics were used to avoid unlikely changes in sleep stages, for example, from awake to deep sleep. Those heuristics included removing short deep sleep stages if they occurred right after an awake period, removing short awake periods if they occurred right after deep periods, and removing all short periods in any of the sleep stages, regardless of the previous sleep stage.

### Data Analyses

First, to determine whether sleep continuity (asleep vs. awake) and sleep architecture (sleep staging) significantly differed between the Happy Ring and both in-laboratory polysomnography and at-home Dreem 2 Headband recordings at time, linear models evaluated mean differences between the values obtained by the Happy Ring and either laboratory PSG or Dreem 2 Headband results, separately. Sleep continuity was evaluated by determining total sleep time and wake after sleep onset. Sleep architecture was defined as time in each of the following sleep stages: deep, light, REM and wake.

Second, classification accuracy was assessed using a confusion matrix, separately evaluating the Happy Generalized and Happy Personalized against both laboratory PSG for the first night of data collection in the lab, and the Dreem 2 Headband for data collected during the two subsequent nights at home. Accuracy was defined as the percent of agreement between each individual investigational device, and the comparison reference (in-lab PSG or at-home Dreem 2 Headband).

Third, accuracy of both the Happy Generalized and Happy Personalized algorithms for sleep versus awake, as well as for individual sleep stages relative to laboratory PSG and Dreem 2 Headband was compared to similar values obtained from Fitbit, Whoop, and Oura devices. Sensitivity, specificity, negative predictive values (NPV) and positive predictive values (PPV) were compared across Happy Generalized, Happy Personalized, Fitbit, Whoop, Oura, and Actiwatch for sleep-wake detection and sleep stage classification separately, using either laboratory PSG or Dreem 2 Headband as the true state for reference.

Fourth, mean differences for TST and WASO, and sleep stages between PSG or Dreem 2 Headband as compared to the wearables (Happy Generalized, Happy Personalized, Fitbit, Whoop, Oura, and Actiwatch) were evaluated for all devices except Actiwatch, which does not provide sleep staging results.

## RESULTS

Data were collected from 40 participants who met inclusion criteria, over the course of 122 nights. After removing data where device failure or data quality issues occurred, data were ultimately analyzed for 36 adults across 77 nights of sleep. The mean age of the sample was 33.8 years (SD=7.8 years, range= 22 years – 51 years). The sample was 56% female, mean body mass index was 23.4 (SD=3.8). The sample was 30.6% non-Hispanic White. A full breakdown of sample characteristics is reported in Table 1.

**Table 1.** Characteristics of the Sample.

| **Variable** | **Category/Units** | **Values** |
| --- | --- | --- |
| **N** | Subjects | N=36 |
| **Sex** | Male | 55.6% |
|  | Female | 44.4% |
| **Age** | Years | 23.4 (SD=3.8) |
| **Body Mass Index** | Kg/m2 | 23.4 (SD=3.8) |
| **Race/Ethnicity** | Non-Hispanic White | 25 (69.4%) |
|  | Hispanic/Latino | 3 (8.3%) |
|  | Asian/Asian American | 6 (16.7%) |
|  | Other/Unknown | 2 (5.6%) |

When polysomnographic values from the laboratory and home references were compared, small, nominal differences were observed. Overall, no significant difference was observed between TST and WASO; mean TST was 432 minutes (SD = 38 minutes) in the lab and 442 minutes (SD = 50 minutes) at home. Mean WASO was 24 minutes (SD = 16 minutes) in the lab and 24 minutes (SD = 14 minutes) at home. The sleep stage distribution was different between lab and home data recordings, with a greater relative percentage of the night in light sleep in the laboratory and greater relative percentages of deep and REM sleep at home. A figure depicting observed values for sleep stages across both in-lab and at-home settings can be found in Supplemental Figure 1. Figure 1 depicts examples of Generalized and Personalized Happy Ring outputs superimposed over in-lab PSG-derived sleep stages and at-home Dreem-derived sleep stages, separately for nights where the Happy Ring performed well, moderately and poorly.

**Figure 1.**
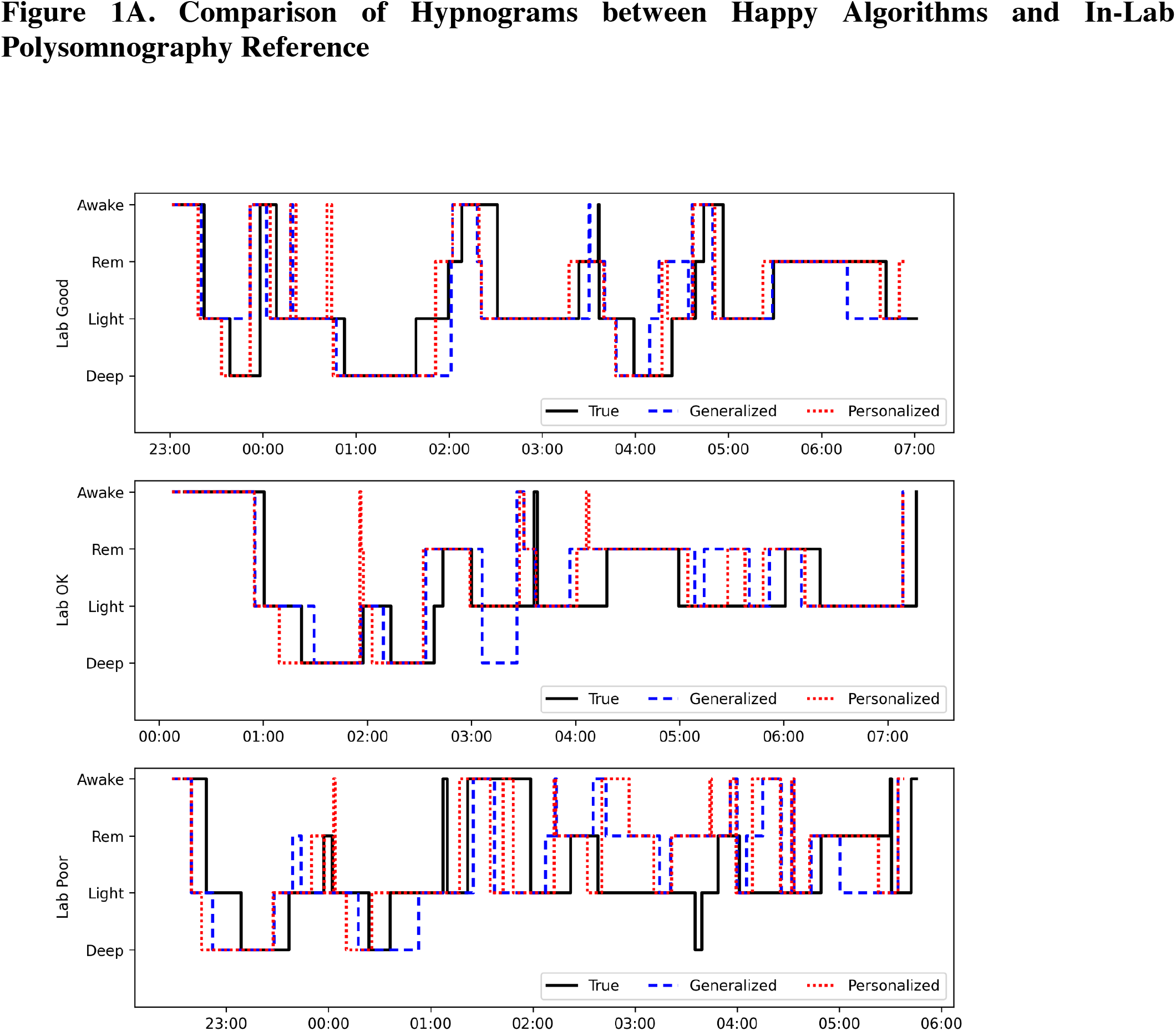
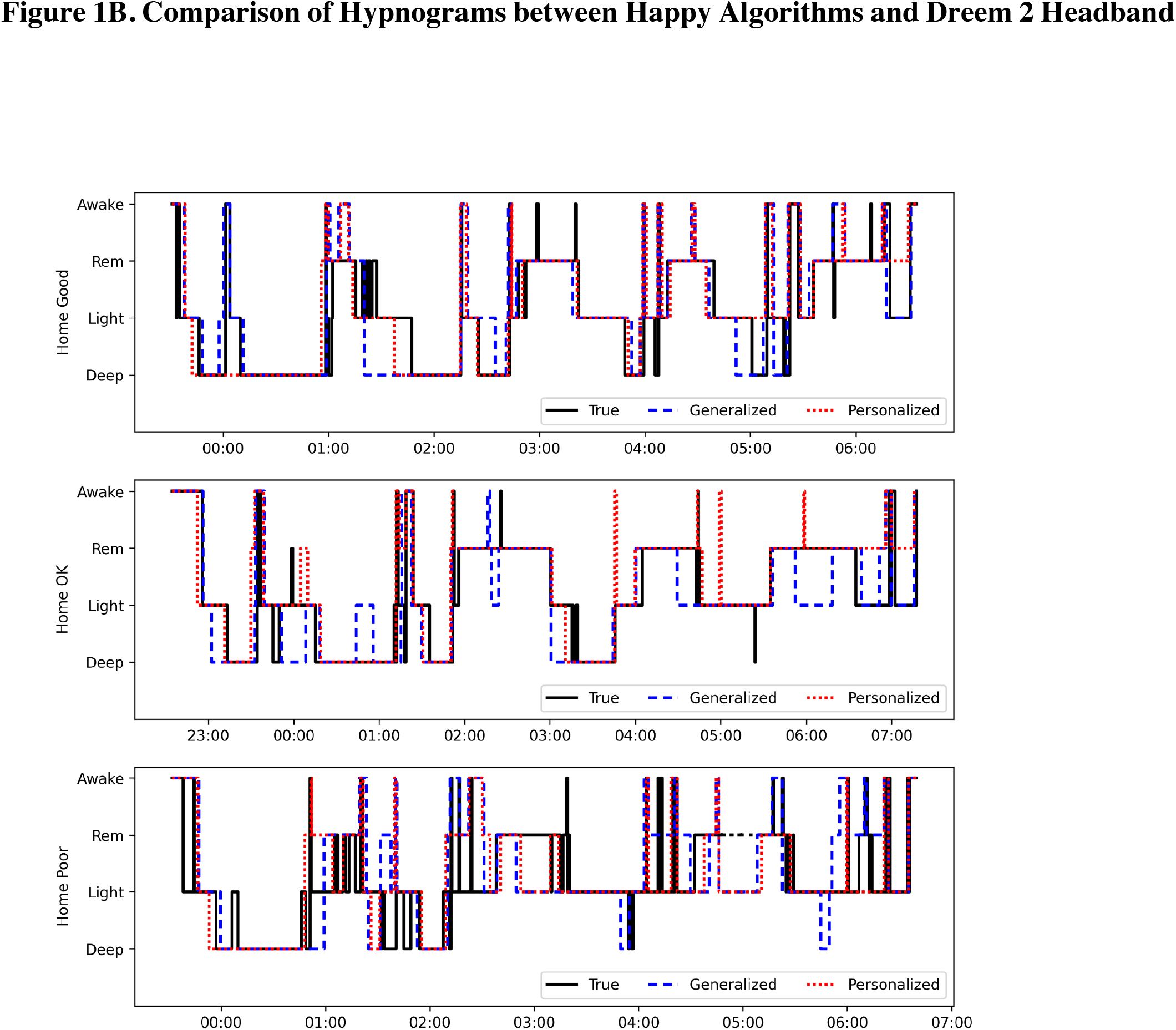
Comparison of Hypnograms between Happy Algorithms and Both In-Lab and At-Home Polysomnography References. In the Good and Typical examples, sleep stages for the Happy Ring display high degrees of overlap with the PSG and Dreem 2 Headband, while the Poor example shows more variation between the two.

### Sleep detection and sleep staging performance in a laboratory setting

Table 2 presents the performance of sleep detection in a laboratory setting for the Happy Personalized and Happy Generalized algorithms, along with that of the other studied devices: Oura, Whoop, Fitbit, and Actiwatch, as compared to the gold standard of laboratory polysomnography. The overall sleep detection accuracy for the Happy devices was 0.92 (SD = 0.04) using the Happy Personalized algorithm, and 0.91 (SD = 0.04) using the Happy Generalized algorithm. Sleep detection accuracy values for other devices fell between 0.84 and 0.88. Table 2 also reports sensitivity, specificity, positive predictive value (PPV) and negative predictive value (NPV) for these devices. Significance of the differences between the Personalized algorithm and the other devices were calculated using paired two-tailed t-tests. In the laboratory environment, the Happy Personalized algorithm demonstrated significantly higher values for accuracy and specificity for sleep detection compared to all other devices studied (*P<*.*001*).

**Table 2.** Sleep Detection of Studied Devices Compared to In-Lab Polysomnography and Dreem 2 Headband At Home.

| *Sleep Detection of Studied Devices Compared to In-Lab Polysomnography* |  |  |  |  |  |  |
| --- | --- | --- | --- | --- | --- | --- |
|  | **Personalized**<br>*Mean ± SD* | **Generalized**<br>*Mean ± SD* | **Oura**<br>*Mean ± SD* | **Whoop**<br>*Mean ± SD* | **Fitbit**<br>*Mean ± SD* | **Actiwatch**<br>*Mean ± SD* |
| **N**<br>*Successful Uses (Distinct Subjects)* | 33 (33) | 33 (33) | 30 (30) | 33 (33) | 27 (27) | 26 (26) |
| **Accuracy** | 0.92±0.04 | 0.91±0.04† | 0.88±0.05† | 0.86±0.05† | 0.84±0.07† | 0.85±0.07† |
| **Sensitivity** | 0.93±0.04 | 0.94±0.04 | 0.93±0.05 | 0.90±0.05* | 0.89±0.09* | 0.94±0.03 |
| **Specificity** | 0.83±0.11 | 0.70±0.18† | 0.49±0.22† | 0.54±0.18† | 0.49±0.18† | 0.19±0.12† |
| **PPV** | 0.97±0.02 | 0.96±0.03† | 0.94±0.04† | 0.93±0.05† | 0.92±0.05† | 0.89±0.07† |
| **NPV** | 0.61±0.23 | 0.61±0.25 | 0.48±0.20† | 0.45±0.21† | 0.39±0.18† | 0.33±0.21† |
| *Sleep Detection of Studied Devices Compared to Dreem 2 Headband* |  |  |  |  |  |  |
| **N**<br>*Successful Uses (Distinct Subjects)* | 40 (26) | 40 (26) | 29 (19) | 31 (20) | 25 (17) | 30 (20) |
| **Accuracy** | 0.92±0.04 | 0.91±0.04† | 0.92±0.03 | 0.89±0.08* | 0.89±0.04* | 0.89±0.03† |
| **Sensitivity** | 0.93±0.04 | 0.94±0.05† | 0.96±0.03* | 0.91±0.09 | 0.92±0.05 | 0.94±0.02 |
| **Specificity** | 0.75±0.11 | 0.67±0.11 | 0.59±0.13† | 0.59±0.21† | 0.60±0.13† | 0.41±0.12† |
| **PPV** | 0.97±0.01 | 0.96±0.03 | 0.96±0.02† | 0.96±0.02† | 0.96±0.03* | 0.93±0.04† |
| **NPV** | 0.54±0.18 | 0.55±0.20† | 0.59±0.18 | 0.49±0.20 | 0.44±0.16† | 0.42±0.17† |

\*indicates significantly different from Personalized $p$ -value $<0.05$
†indicates significantly different from Personalized $p$ -value $<0.001$

The performance of sleep stage predictions for light, deep, REM and wake were also evaluated. For light, deep, and REM sleep, Happy Personalized demonstrated comparable values for sensitivity, specificity, and accuracy to other devices studied. Values for accuracy, sensitivity, specificity, PPV and NPV for all study devices compared to the lab PSG can be found in Supplemental Table 1. For data collected in the sleep laboratory, total sleep time estimates, WASO, and sleep stages, relative to PSG values, are depicted in Figure 2A, 2B, and 2C, respectively for each of the devices studied.

**Figure 2.**
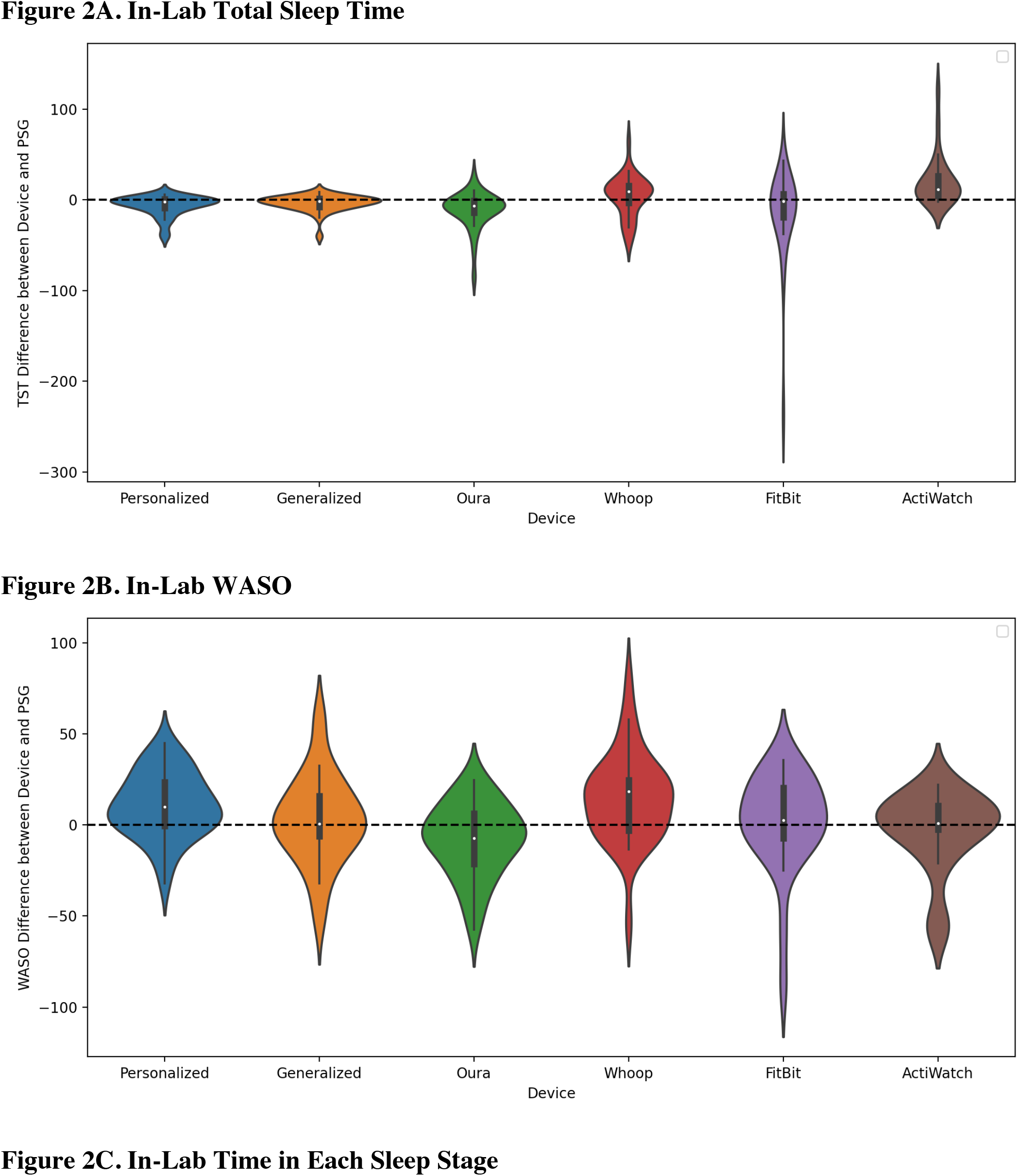
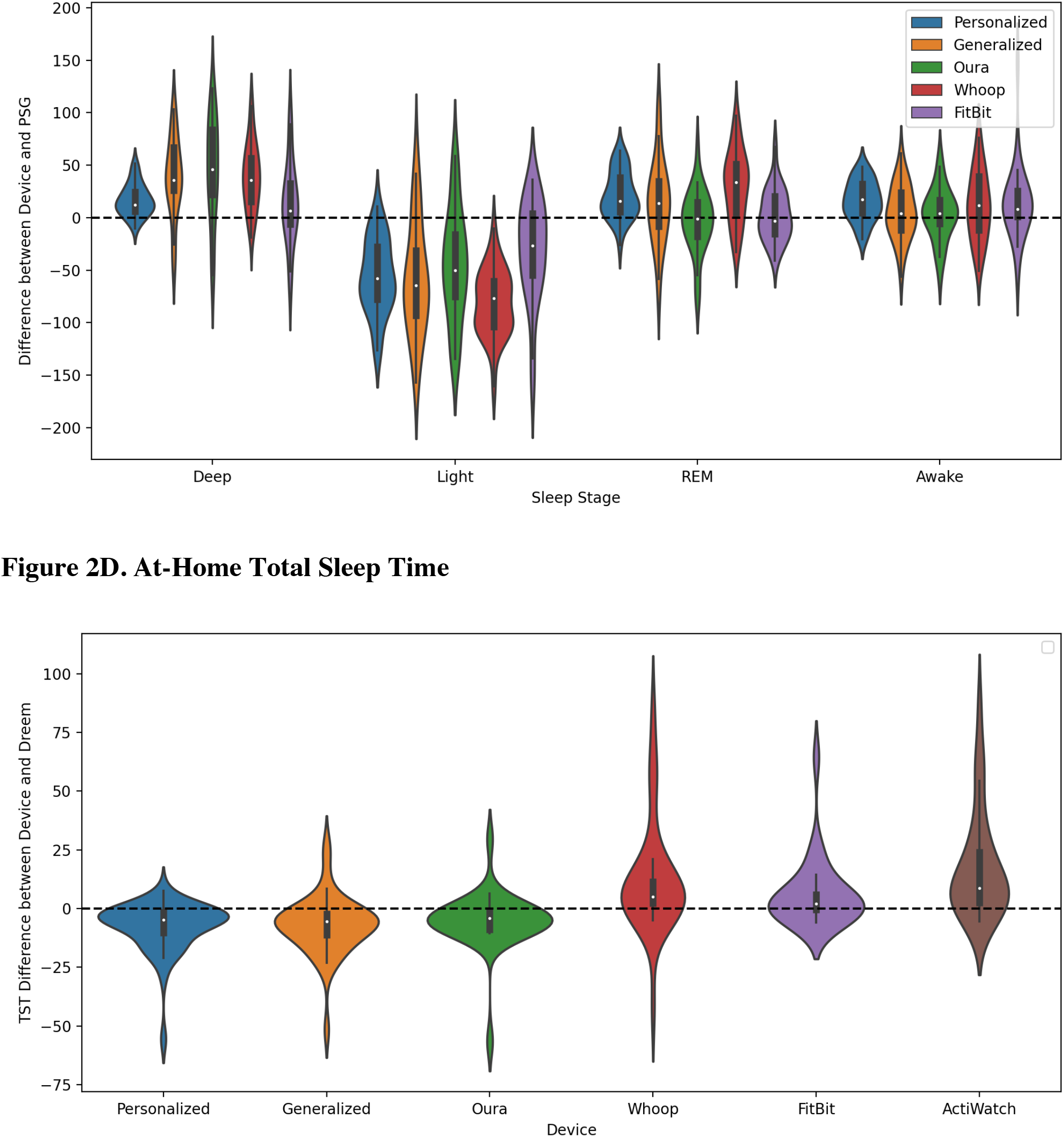
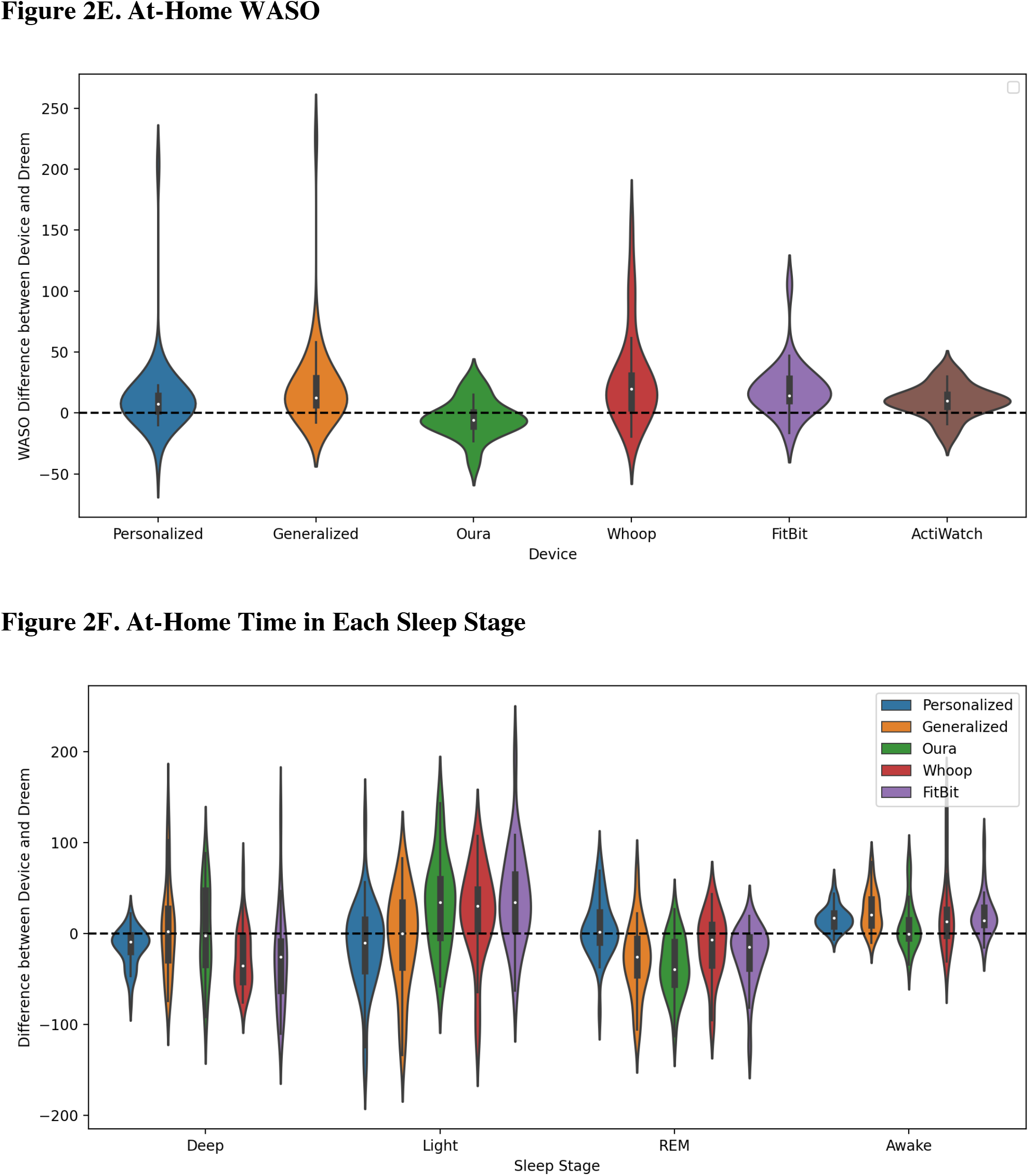
Comparison of Deviations from In-Lab and At-Home Polysomnography for Happy Algorithms and Other Studied Devices. Violin plots demonstrate the aggregation of data near to the line of no difference in measures, with positive values signifying over detection by the Happy Ring and negative values signifying under detection. The width of the plot signifies the density of values near a given value, and the tails extend to the full ranges of the outputs. The gray box plot inside the violin plot plots the median, interquartile range, and data range excluding outliers.

Bland-Altman plots comparing the Happy Generalized and Happy Personalized algorithms were computed. Data comparing these algorithms relative to sleep continuity and sleep stages are reported in Figure 3A and Figure 3B, presenting Happy Generalized and Happy Personalized, respectively.

**Figure 3.**
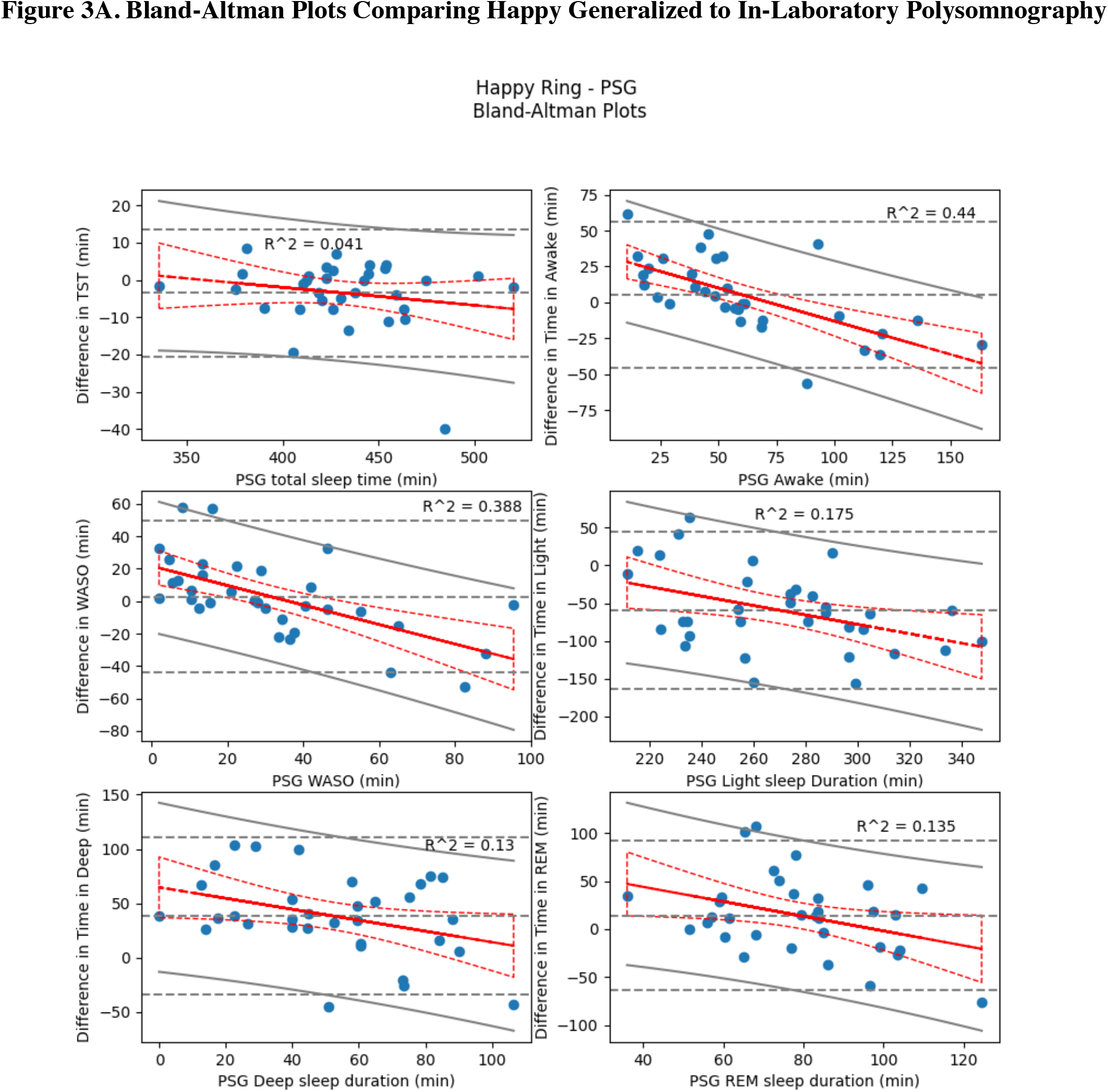
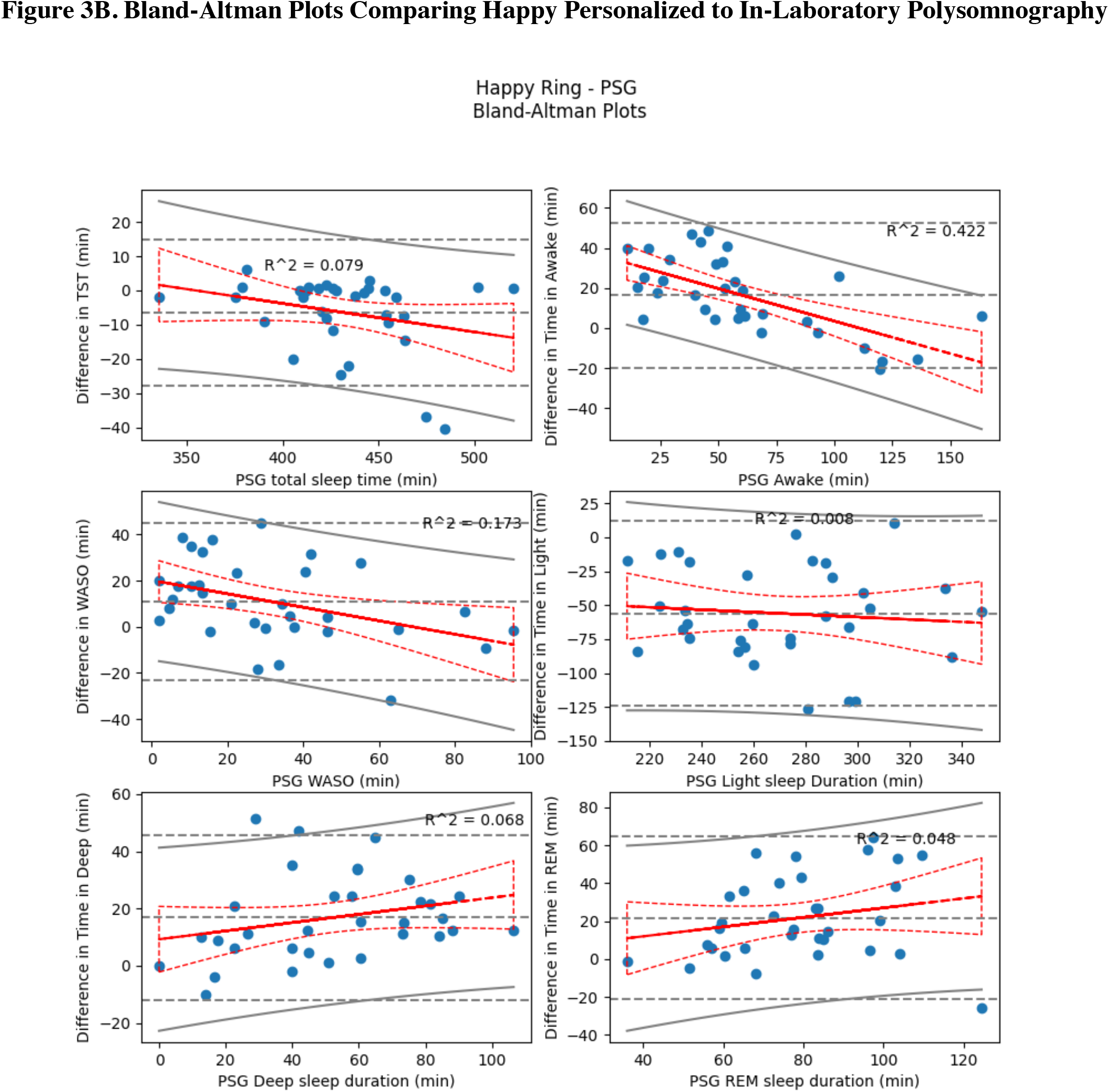
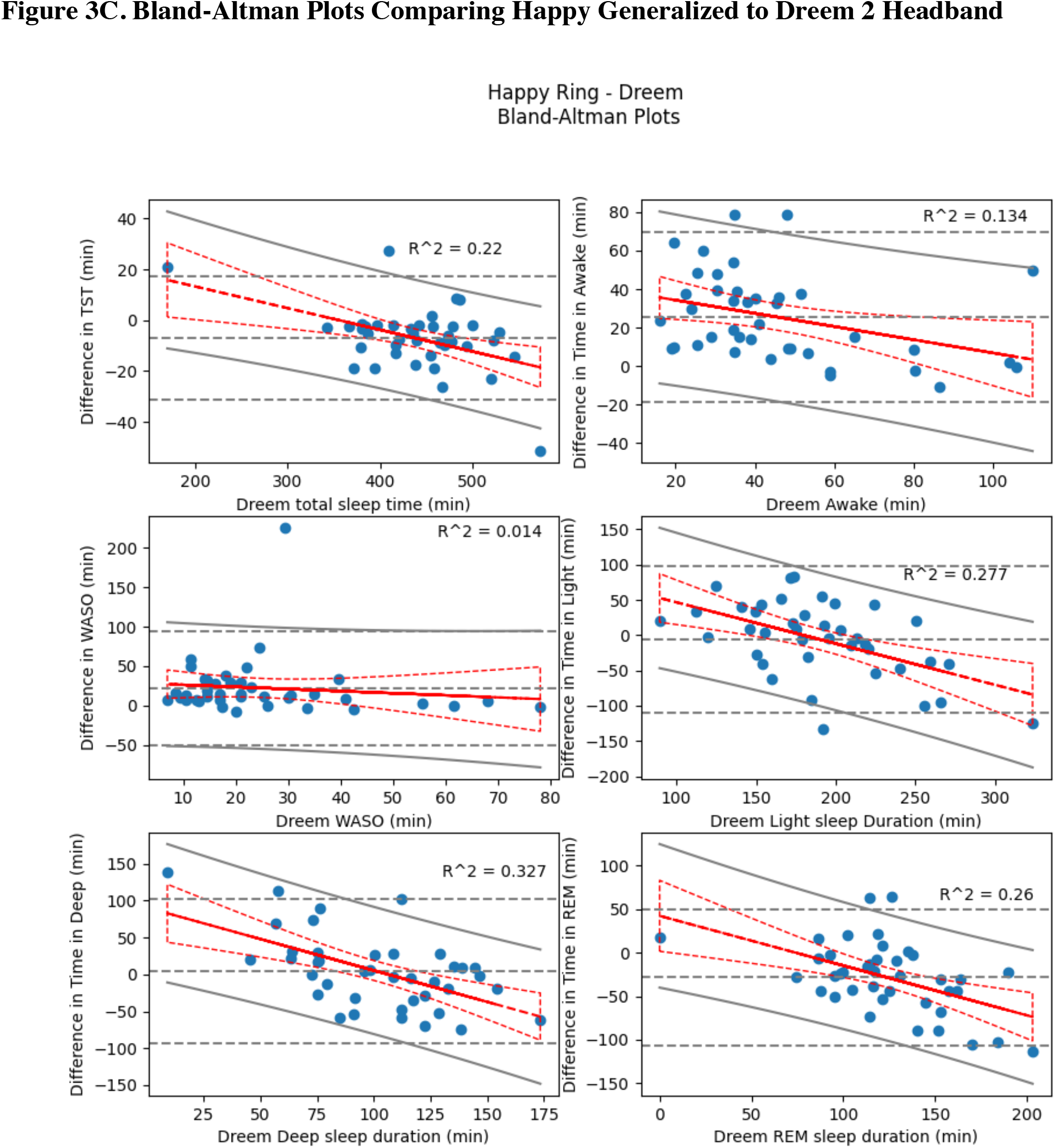
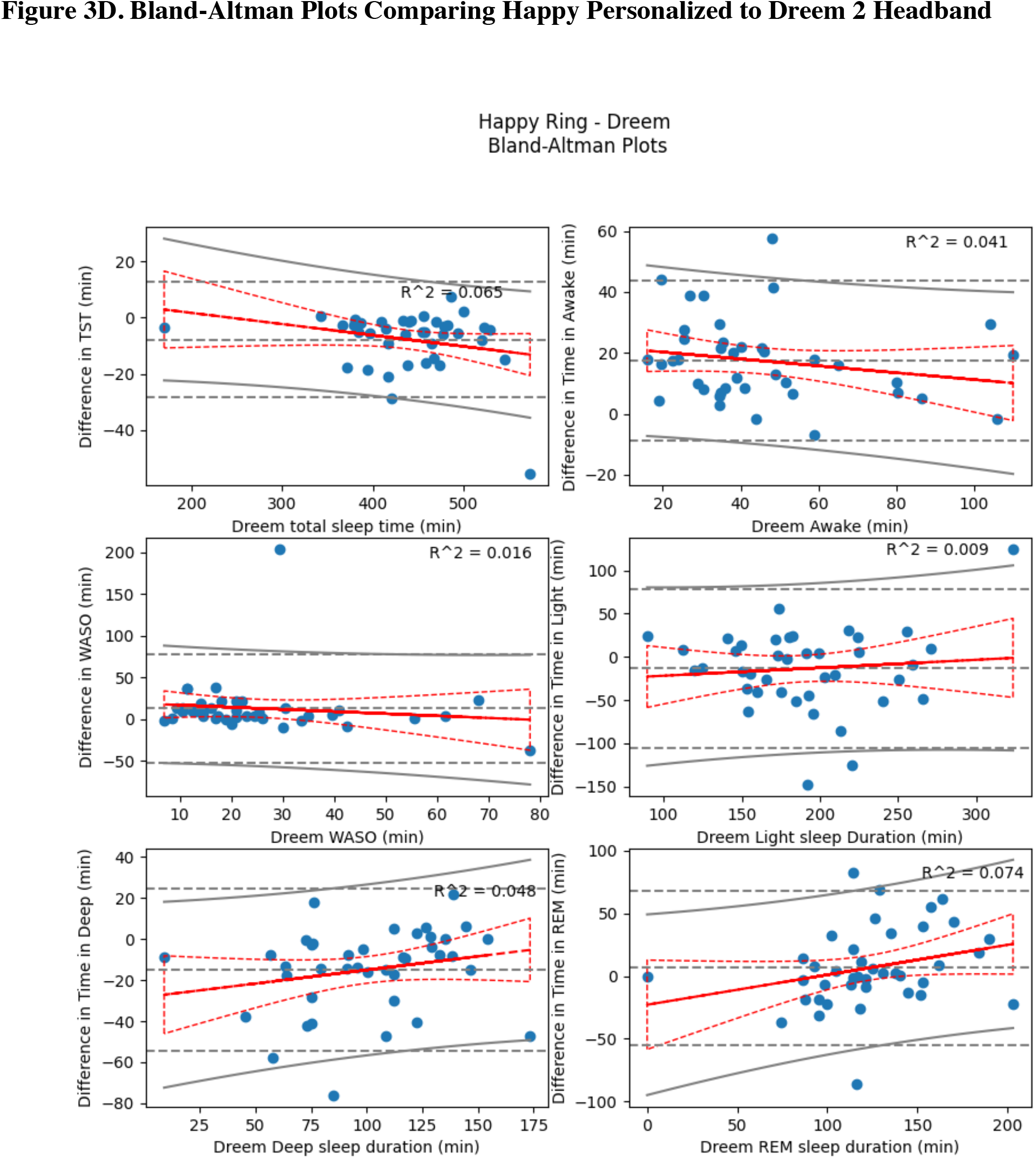
Bland-Altman Plots Comparing Happy Generalized to In-Laboratory Polysomnography and Dreem 2 Headband.

### Sleep detection and sleep-staging performance in a home setting

The sleep detection performance at-home of the Happy Personalized and Happy Generalized algorithms, along with those of Oura, Whoop, Fitbit, and Actiwatch devices are presented in Table 2. The overall accuracy for the Happy devices was 0.92 (SD = 0.04) for the Happy Personalized algorithm and 0.91 (SD = 0.04) for the Happy Generalized algorithm when compared to the Dreem 2 Headband. Values for sleep detection accuracy at home for other devices feel between 0.89 and 0.92. Specificity, sensitivity, positive predictive value and negative predictive values for sleep detection for these devices are also reported in Table 2.

For data collected at home, TST estimates, relative to Dreem 2 Headband values, are depicted in Figure 2D, and differences in WASO across devices are reported in Figure 2E. Figure 2F shows differences for sleep stages across devices. Bland-Altman plots comparing the Happy Generalized and Happy Personalized algorithms were computed. Data comparing these algorithms relative to sleep continuity and sleep stages are reported in Figure 3C and Figure 3D.

### Confusion Matrices

Confusion matrices are depicted in Figure 4. Figure 4A depicts the 4-stage confusion matrix comparing Happy Generalized and in-lab polysomnography. Figure 4B depicts the confusion matrix for Happy Personalized and in-lab polysomnography. Figures 4C and 4D depict the confusion matrices for Happy Generalized and Happy Personalized at home, respectively. Due to unbalanced classes, values presented are each prediction’s normalized detection frequency with regards to the reference (PSG or Dreem 2 Headband), including mean, standard deviation, and confidence interval. The color indicates the scaled probability of occurrence of a specific pair, hence artificially removing class imbalance. The title in each plot shows the average classification accuracy across all sleep stages.

**Figure 4.**
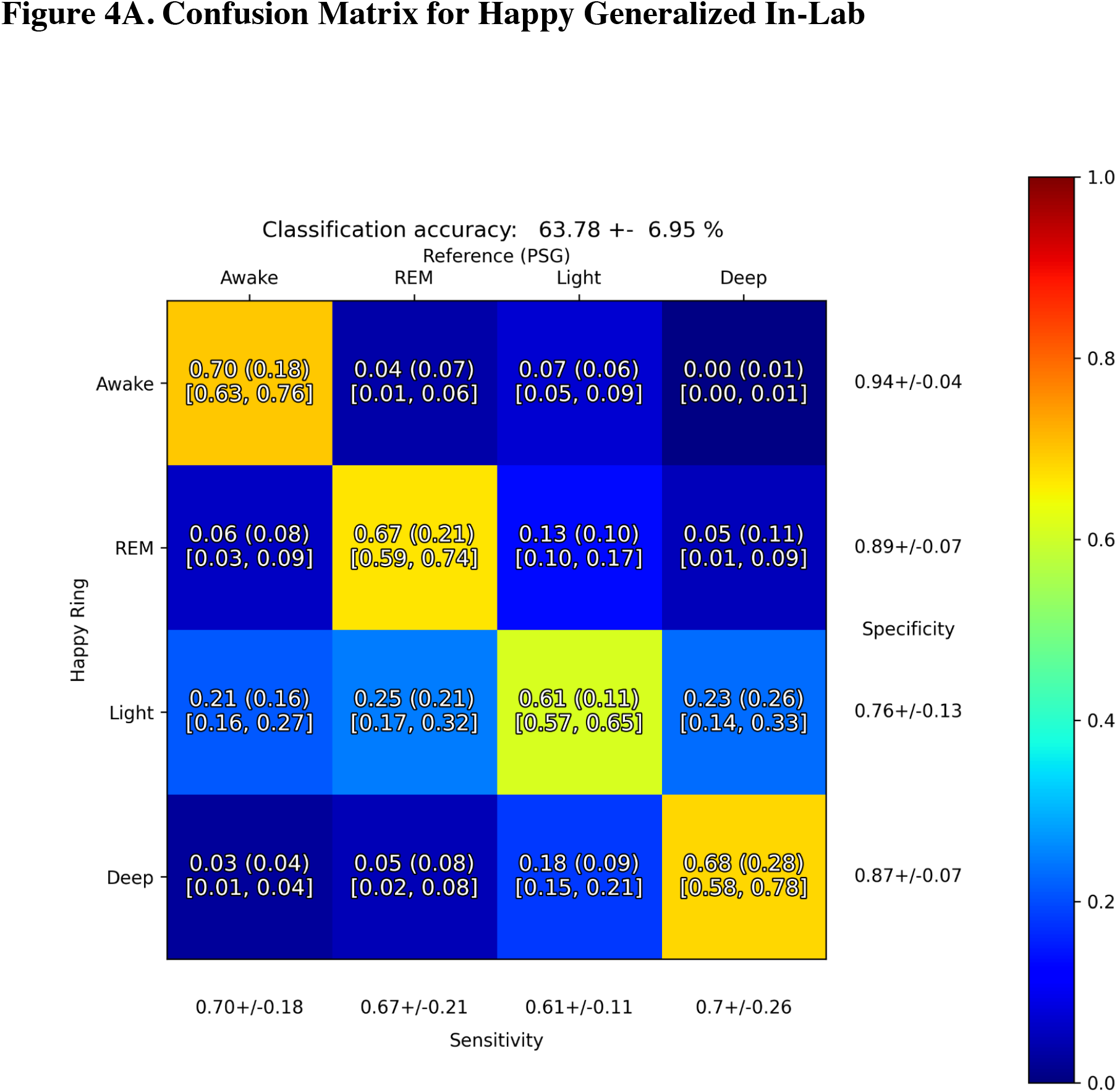
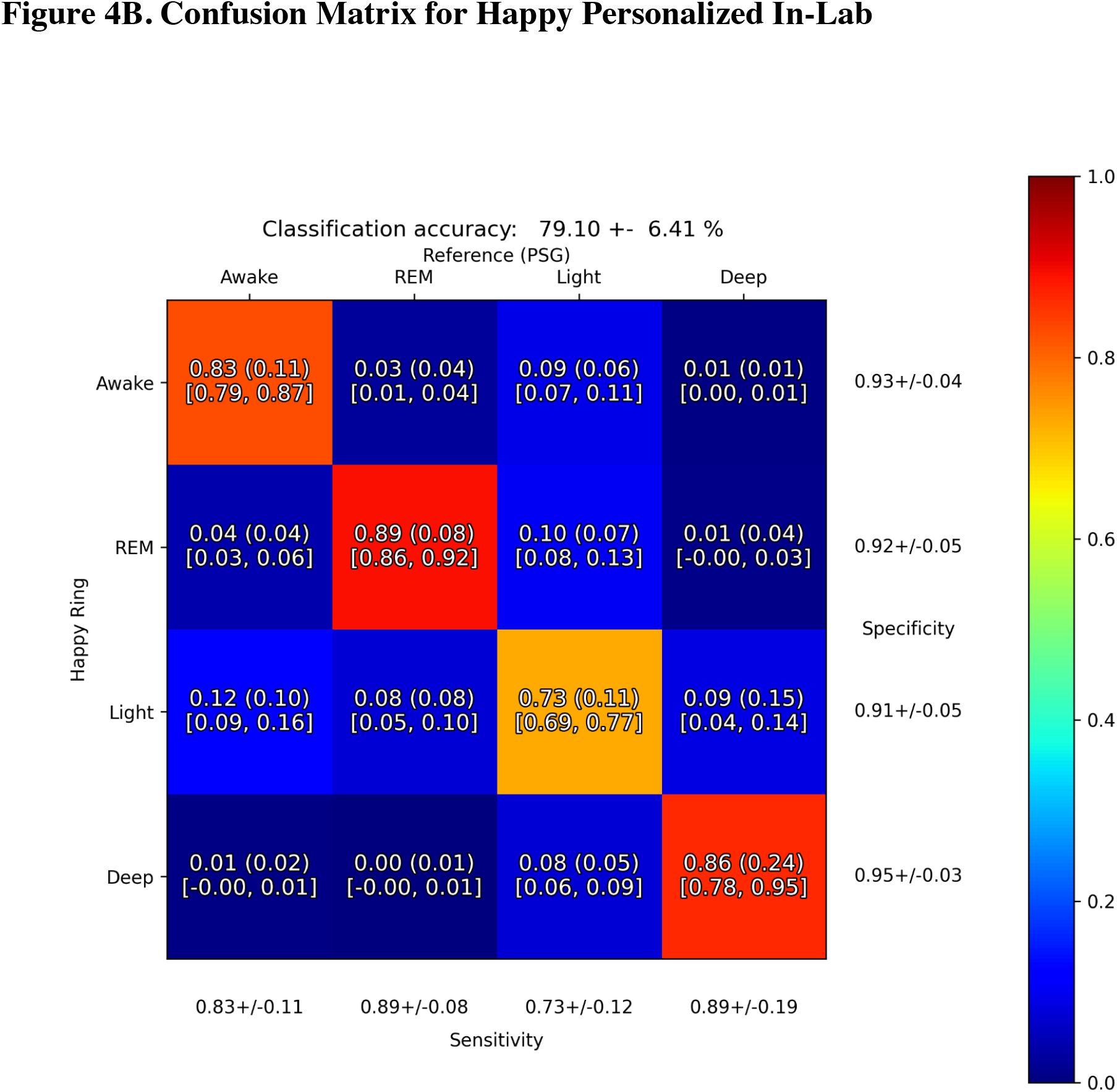
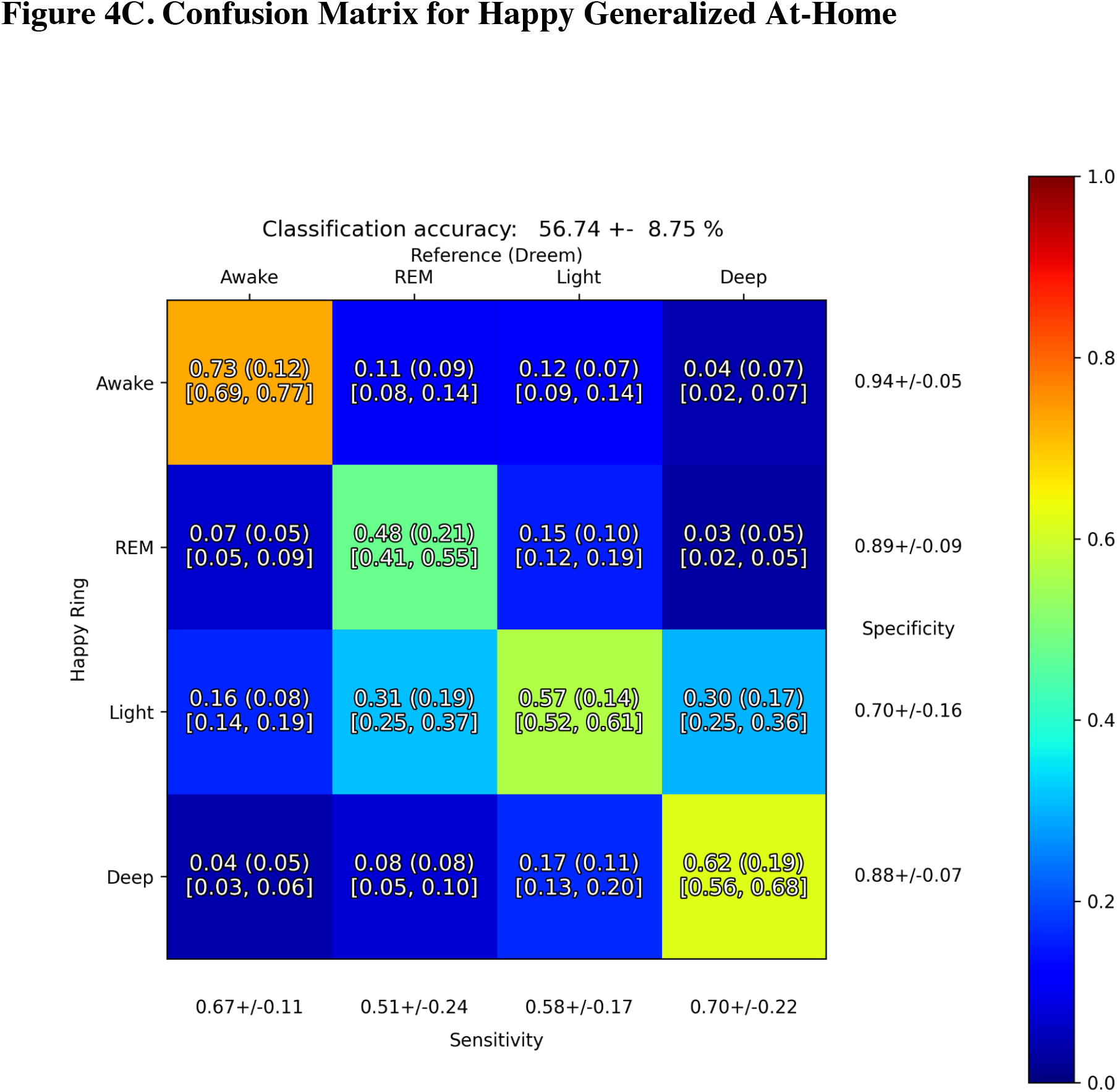
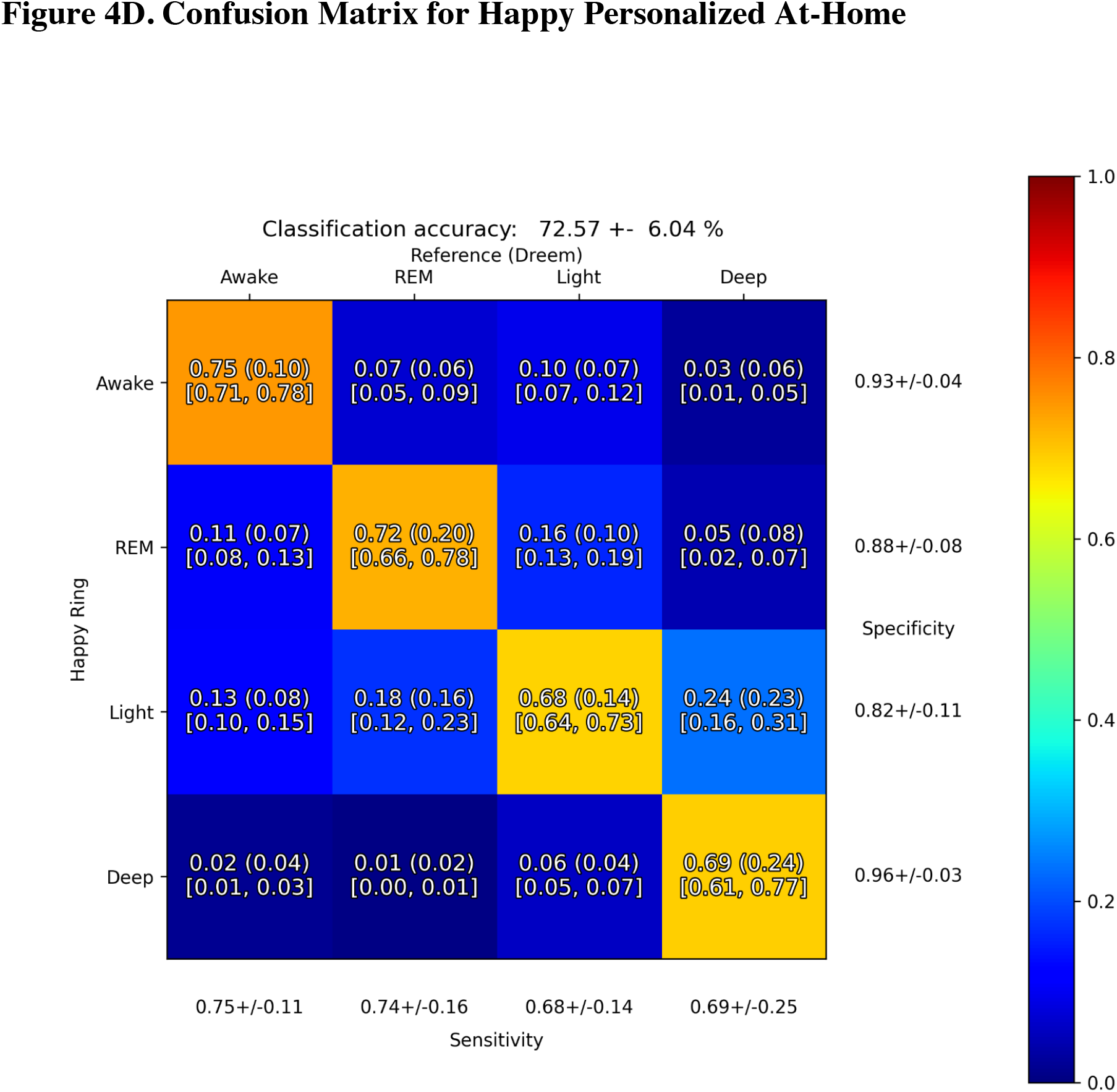
Confusion Matrices Comparing Happy Algorithms to In-Lab Polysomnography and Dreem 2 Headband References. Each box includes the Mean Accuracy and the (Standard Deviation) and [95% Confidence Interval].

## DISCUSSION

The present study evaluated the performance of the Happy Ring for the evaluation of sleep continuity and sleep architecture among healthy working-age adults, using two different scoring approaches – Generalized and Personalized. Compared to both laboratory polysomnography and at-home Dreem 2 Headband, the Generalized scoring strategy demonstrated sensitivity and specificity for sleep-wake detection similar to other comparable devices that have been previously empirically evaluated. The Personalized scoring strategy maintained similar sensitivity to the Generalized approach, but also demonstrated improved specificity relative to the other devices studied for sleep-wake detection (*P<*.*001*). Regarding sleep staging, the rate of agreement between the Happy Ring and both the Dreem 2 Headband and laboratory polysomnography for light, deep, and REM sleep was moderate, similar to other sleep-tracking devices to which it was compared. The Personalized scoring approach, again, demonstrated improved accuracy for discernment of sleep stages compared to other devices evaluated (*P< 0*.*001*). Taken together, the Happy Ring and associated sleep scoring algorithms demonstrated good performance for the detection of sleep versus wake, relative to both home and laboratory references, in line with other devices that also use movement and heart rate to evaluate sleep. Further, the Personalized scoring approach consistently performed better than the Generalized approach.

The main finding from this work is that, among generally healthy adults, the Happy Ring performs comparably in terms of discerning sleep versus wake when compared to other similar devices in both a lab and an at-home setting. The degree of sensitivity (93-94%) was similar to the other devices that were evaluated (89-94%). This is in line with previous work, which shows that these and similar devices typically demonstrate sensitivity over 90%^28,45^. However, it should be noted that the sensitivity found for some of the comparison devices in this study were a few percentage points below that seen in previously published studies using those same devices. This finding could be a result of a myriad of factors, such as a more diverse sample in the present study, the presence of an at-home component of the protocol, or may be related to other unknown factors.

When evaluating sleep detection, specificity refers to the ability of a device to detect wakefulness in the context of otherwise continuous sleep. This is the parameter for which current devices tend to perform less well. A recent paper by Chinoy and colleagues compared 7 different devices and found specificity values between 18-54%^28^. These findings are similar to those observed in the present study, which found specificity values ranging from 19-54% across previously-studied devices relative to in-laboratory polysomnography. The Happy Generalized algorithm performed nominally better, with 70% specificity in-lab. The Happy Personalized approach demonstrated statistically significantly improved performance over the other devices evaluated, with a mean specificity of 83% in-lab and 75% at home (*P< 0*.*001*). This suggests that the personalized approach may be a strategy for improving sleep detection by improving the ability of the device to discern wake epochs. This is especially important for the evaluation of relatively healthy sleepers, as most healthy sleepers will achieve a sleep efficiency of over 90%^48^. If a device simply scores every epoch as sleep, with no discernment or algorithm, it will appear to have 90% accuracy. In this case, it would have 100% sensitivity but 0% specificity. Therefore, for a consumer device that will presumably be used to assess sleep in individuals with generally high sleep efficiency, higher specificity is perhaps more important than overall accuracy because of the skewed nature of sleep-wake data in healthy populations. For this reason, the improved specificity of the Happy Personalized scoring approach may represent a significant innovation that can improve the utility of wearable sleep assessments.

Regarding sleep stages, the Happy Ring sleep scoring algorithms performed relatively well at discerning light (stages N1 and N2), deep (stage N3) and REM sleep. Previous studies have shown that peripheral data from heart rate and movement can be used to approximate sleep stages, but these algorithms are still limited in providing more than a general approximation^30,49^. For this reason, peripherally scored sleep staging data are still insufficient for replacing sleep stages derived from polysomnography. Regardless, values from at-home sleep tracking devices can be useful, especially as they can be sampled over many days and in large numbers of people, unlike polysomnography^9,11^. For sleep stage classification, the performance of the Happy Generalized model performed about the same as other similar devices on the market both in-lab and at-home. More importantly, the Happy Personalized model performed significantly better than other comparable devices for classification of all sleep stages in regards to accuracy, sensitivity, PPV, and NPV in the lab (*P < 0*.*001*).

We also evaluated the level of agreement between the Happy Ring (Personalized and Generalized) and in-laboratory PSG as well as the Dreem 2 Headband via discrepancy analysis, graphically displayed as Bland-Altman plots (Figure 3). Bland-Altman plots are considered to be a key method in analyzing the agreement between two continuous medical measurements^10^. The majority of measures showed a negative proportional bias, whose magnitude depended on the range of PSG or Dreem 2 Headband measures. In other words, sleep continuity and staging tended to be underestimated by the Happy Ring for cases showing higher PSG-derived or Dreem 2 Headband-derived measures. This was not always the case, however, as some measures showed very little bias. Such was the case with WASO and light sleep derived from the Happy Ring relative to the Dreem 2 Headband, indicating close agreement. In general, the Happy Personalized model demonstrated slightly better performance than the Generalized model, reflected by an overall lower bias and more narrow limits of agreement across measures. Thus, the Happy Ring compares relatively well to both PSG and the Dreem 2 Headband, although discrepancies of sleep variables tended to be larger as “true” measures of sleep increased.

The first actigraphy scoring studies used hand scoring^50^. Soon afterwards, scoring algorithms were introduced^51^ and subsequently refined^52,53^. These algorithms started as basic prediction equations that used weighted values from the epoch of interest and multiple epochs in the past or future to determine, based on movement, if an individual is awake or asleep^52,53,54^. Over time, and with the inclusion of other signals like heart rate, these algorithms have become more complex^28,41^. A more recent advance is the use of machine learning to not only derive an optimal scoring algorithm based on a specific signal, but also as part of a strategy to combine multiple signals^4,32,33^. Although this approach has demonstrated improvements over older approaches, there is still the limitation that the algorithm itself remains largely static and generalized to the population, rather than to the individual. However, with the advent of improved computing power, it is possible to develop an algorithm that is dynamic and changes based on the users’ own data. This personalization is the difference between the Happy Generalized and Personalized approaches. The Generalized approach, as evaluated in this study, represents a relatively static algorithm that is universally applied to all records, which is the standard approach across other devices. Since a generalized approach is functionally similar to the algorithms in the other studied devices, it is unsurprising that this algorithm performs comparably well. The Personalized scoring approach, however, represents a potential innovation, in that it starts with generalized parameters but changes over time based on use. As the user wears the device over multiple subsequent recording periods, the algorithm modifies itself in response to the parameters obtained by that user. Thus, the scoring algorithm is fluid and unique across users, based on their own data. This approach – a dynamic, personalized algorithm – demonstrated the best performance both in the lab and at home, especially for the key parameter of specificity. Future algorithm development for sleep wearables may be improved by including an individualized scoring component.

This study had several unique strengths. Objective polysomnographic sleep was assessed both in the laboratory and at home, under naturalistic conditions. Further, the performance of the Happy Ring was assessed relative to other types of devices, including standard actigraphy (i.e., Actiwatch), wrist-worn multisensory device (i.e., Fitbit, Whoop), and ring-worn multisensory device (i.e., Oura). The design of the study and the subsequent analytic methods were aligned with recent guidelines and recommendations for sleep-tracking device evaluation^9,10^.

However, the study did have some limitations. The sample size was relatively small and consisted of generally healthy adults, which may limit generalizability of the findings. Future research in evaluation of the Happy Ring and associated sleep algorithms will aim to collect data from a wider, more diverse population. Additionally, the home-based comparison recordings were obtained using a commercial device (i.e., Dreem 2 Headband) rather than traditional laboratory polysomnography. Lastly, the algorithms underlying the Happy Generalized and Happy Personalized scoring strategies are considered proprietary and rely on undisclosed factors.

In conclusion, the present study describes the initial development and assessment of the Happy Ring, a novel device for the assessment of sleep continuity and sleep architecture. Overall, the device performed well, demonstrating good sensitivity and moderate specificity, comparable to other consumer devices on the market. When compared to other consumer sleep-tracking devices, the Happy Generalized scoring procedure demonstrated similar performance for sleep detection and sleep stage classification; while the novel Happy Personalized algorithm demonstrated significant improvement in performance compared to other devices studied.

Future research should examine the performance of this device in other contexts and larger samples. In addition, future work should explore the value of additional peripheral signals to the assessment of sleep in the context of overall health and wellbeing. Finally, the improved performance of the Happy Personalized algorithm suggests that sleep assessment using wearables can be improved by developing more dynamic and adaptable algorithms that leverage user data prospectively.

## Data Availability

Data from this study are not available due to the raw data being important intellectual property for Happy Health, Inc.

## DISCLOSURES

Financial conflict of interest disclosures: Zohar Bromberg, Aaron Hadley, Zoe Morrell, Arnulf Graf and Dustin Freckleton are employees of Happy Health, Inc.

Non-financial conflict of interest disclosures: Michael Grander reports grants from Jazz Pharmaceuticals and CeraZ, and has received consulting fees in the past 24 months from Fitbit, Natrol, Casper, Athleta, Smartypants Vitamins, Idorsia, Jazz Pharmaceuticals, New York University, and University of Maryland.

Data collection and data analyses were performed by members of the Happy Health, Inc. team listed as authors in this paper.

Authors listed as affiliated with the University of Arizona assisted with study design, data analysis interpretation, and preparation for publication.

## Notes

### Competing Interest Statement

Zohar Bromberg, Aaron Hadley, Zoe Morrell, Arnulf Graf and Dustin Freckleton are employees of Happy Health, Inc. Michael Grander reports grants from Jazz Pharmaceuticals and CeraZ, and has re-ceived consulting fees in the past 24 months from Fitbit, Natrol, Casper, Athleta, Smartypants Vit-amins, Idorsia, Jazz Pharmaceuticals, New York University, and University of Maryland. Stephen Hutchinson reports no potential conflicts.

### Funding Statement

This study did not receive any funding.

### Author Declarations

Solutions Institutional Review Board gave ethical approval for this work.

### Summary of Updates

An additional author was added to the manuscript (Aaron Hadley). The manuscript has been revised for clarity, including additional information on statistical significance in differences between devices in an effort to strengthen the validity of the findings. Tables 2 + 4 were combined, and Tables 3 + 5 were moved to Supplemental Materials. Figure 1 was pared down and moved to Supplemental Materials, Figure 2 was pared down, Figures 3 +4 were modified for clarity.

