## Supplemental Materials for "Performance of a Multisensor Smart Ring to Evaluate Sleep: In-Lab and Home-Based Evaluation Relative to Polysomnography and Actigraphy: Importance of Generalized Versus Personalized Scoring"

Short running title: *Performance of a Multisensor Smart Ring to Evaluate Sleep*

Data Accessibility statement: Data from this study are not available due to the raw data being important intellectual property for Happy Health, Inc.

Keywords: sleep technology, wearables, validation, polysomnography, actigraphy, sensors

**Supplemental Figure 1. Comparison between Laboratory PSG and At-Home Dreem Recording.** Bar plots represent the mean with standard deviation, and there were significant differences (* $p<0.05$) between Lab and Home for Deep, Light, and REM stage detection.

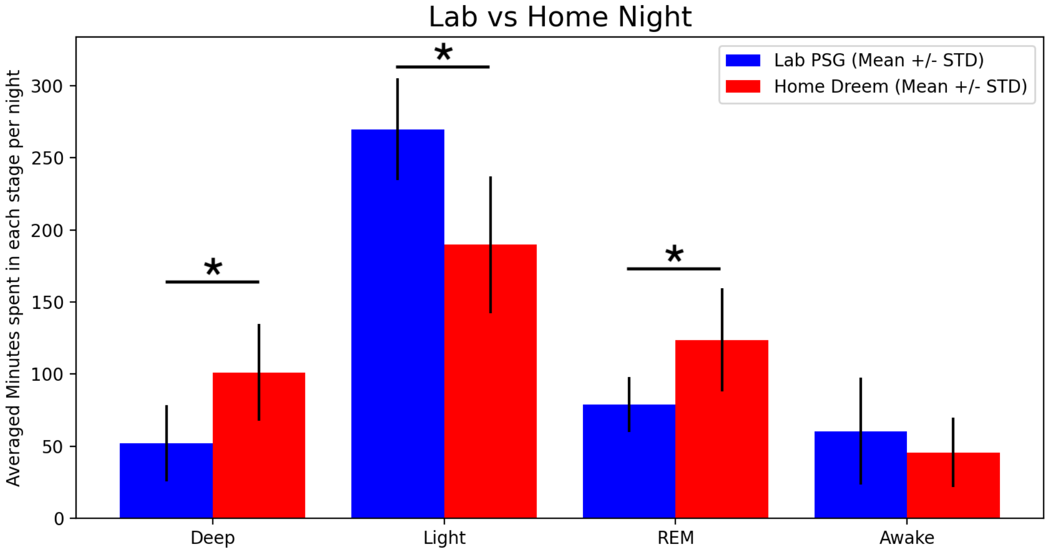

Lab vs Home Night
Lab PSG (Mean +/- STD)
Home Dreem (Mean +/- STD)
Averaged Minutes spent in each stage per night
0
50
100
150
200
250
300
Deep
Light
REM
Awake
*
*
*

**Supplemental Table 1. Sleep Stage Detection with Studied Devices Compared to In-Lab Polysomnography**

| | **Personalized** *Mean ± SD* | **Generalized** *Mean ± SD* | **Oura** *Mean ± SD* | **Whoop** *Mean ± SD* | **Fitbit** *Mean ± SD* |
|---|---|---|---|---|---|
| **N** *Total Recordings (Unique Subjects)* | 33 (33) | 33 (33) | 30 (30) | 33 (33) | 27 (27) |
| **Awake** | | | | | |
| **Accuracy** | 0.92±0.04 | 0.91±0.04 | 0.88±0.05† | 0.86±0.05† | 0.84±0.07† |
| **Sensitivity** | 0.83±0.11 | 0.70±0.18† | 0.49±0.22† | 0.54±0.18† | 0.49±0.18† |
| **Specificity** | 0.93±0.04 | 0.94±0.04 | 0.93±0.05 | 0.90±0.05* | 0.89±0.09* |
| **PPV** | 0.61±0.23 | 0.61±0.25 | 0.48±0.20† | 0.45±0.21† | 0.39±0.18† |
| **NPV** | 0.97±0.02 | 0.96±0.03† | 0.94±0.04† | 0.93±0.05† | 0.92±0.05† |
| **Light** | | | | | |
| **Accuracy** | 0.81±0.06 | 0.67±0.07† | 0.61±0.09† | 0.59±0.06† | 0.65±0.07† |
| **Sensitivity** | 0.73±0.12 | 0.61±0.11† | 0.57±0.14† | 0.49±0.09† | 0.64±0.12* |
| **Specificity** | 0.91±0.05 | 0.76±0.13† | 0.64±0.13† | 0.71±0.10† | 0.66±0.13† |
| **PPV** | 0.92±0.05 | 0.79±0.10† | 0.69±0.08† | 0.71±0.08† | 0.73±0.07† |
| **NPV** | 0.71±0.10 | 0.58±0.10† | 0.53±0.12† | 0.50±0.08† | 0.57±0.11† |
| **Deep** | | | | | |
| **Accuracy** | 0.95±0.03 | 0.85±0.06† | 0.81±0.08† | 0.82±0.05† | 0.86±0.04† |
| **Sensitivity** | 0.89±0.19 | 0.70±0.26† | 0.60±0.26† | 0.56±0.19† | 0.48±0.26† |
| **Specificity** | 0.95±0.03 | 0.87±0.07† | 0.83±0.10† | 0.85±0.05† | 0.91±0.05* |
| **PPV** | 0.67±0.18 | 0.38±0.22† | 0.35±0.17† | 0.34±0.20† | 0.40±0.30† |
| **NPV** | 0.99±0.01 | 0.96±0.04† | 0.94±0.04† | 0.94±0.04† | 0.94±0.04† |
| **REM** | | | | | |
| **Accuracy** | 0.92±0.04 | 0.85±0.05† | 0.84±0.05† | 0.81±0.07† | 0.86±0.05† |
| **Sensitivity** | 0.89±0.08 | 0.67±0.21† | 0.51±0.21† | 0.65±0.19† | 0.61±0.17† |
| **Specificity** | 0.92±0.05 | 0.89±0.07* | 0.91±0.05 | 0.85±0.08† | 0.91±0.05 |
| **PPV** | 0.72±0.12 | 0.58±0.17† | 0.55±0.14† | 0.49±0.18† | 0.60±0.16† |
| **NPV** | 0.98±0.02 | 0.93±0.05† | 0.90±0.04† | 0.92±0.04† | 0.92±0.04† |
| **All Stages** | | | | | |
| **Sensitivity** | 0.80±0.07 | 0.64±0.07† | 0.57±0.09† | 0.54±0.08† | 0.61±0.07† |
| **Kappa** | 0.68±0.09 | 0.45±0.11† | 0.32±0.13† | 0.32±0.10† | 0.37±0.13† |

**indicates significantly different from Personalized p-value <0.05*
*†indicates significantly different from Personalized p-value <0.001*

**Supplemental Table 2. Sleep Stage with Studied Devices Compared to Dreem 2 Headband**

| | **Personalized** *Mean ± SD* | **Generalized** *Mean ± SD* | **Oura** *Mean ± SD* | **Whoop** *Mean ± SD* | **Fitbit** *Mean ± SD* |
|---|---|---|---|---|---|
| **N** *Total Recordings (Unique Subjects)* | 40 (26) | 40 (26) | 29 (19) | 31 (20) | 25 (17) |
| **Awake** | | | | | |
| **Accuracy** | 0.92±0.04 | 0.91±0.04 | 0.92±0.03 | 0.89±0.08* | 0.89±0.04* |
| **Sensitivity** | 0.75±0.11 | 0.67±0.11 | 0.59±0.13 | 0.59±0.21 | 0.6±0.13 |
| **Specificity** | 0.93±0.04 | 0.94±0.05† | 0.96±0.03* | 0.91±0.09 | 0.92±0.05 |
| **PPV** | 0.54±0.18 | 0.55±0.20† | 0.59±0.18 | 0.49±0.20 | 0.44±0.16† |
| **NPV** | 0.97±0.01 | 0.96±0.03 | 0.96±0.02† | 0.96±0.02† | 0.96±0.03† |
| **Light** | | | | | |
| **Accuracy** | 0.78±0.05 | 0.66±0.08† | 0.67±0.07† | 0.67±0.09† | 0.7±0.10† |
| **Sensitivity** | 0.68±0.14 | 0.58±0.17† | 0.68±0.13 | 0.66±0.15 | 0.73±0.11 |
| **Specificity** | 0.82±0.11 | 0.70±0.16† | 0.65±0.14† | 0.69±0.11† | 0.67±0.13† |
| **PPV** | 0.75±0.11 | 0.60±0.13† | 0.58±0.10† | 0.58±0.12† | 0.62±0.12† |
| **NPV** | 0.80±0.06 | 0.72±0.11† | 0.75±0.08* | 0.76±0.09 | 0.78±0.10 |
| **Deep** | | | | | |
| **Accuracy** | 0.92±0.03 | 0.83±0.05† | 0.84±0.05† | 0.86±0.06† | 0.84±0.08† |
| **Sensitivity** | 0.69±0.25 | 0.70±0.22 | 0.63±0.32* | 0.59±0.17 | 0.56±0.26* |
| **Specificity** | 0.96±0.03 | 0.88±0.07† | 0.89±0.08† | 0.95±0.04 | 0.94±0.06 |
| **PPV** | 0.85±0.12 | 0.61±0.21† | 0.64±0.19† | 0.78±0.15 | 0.71±0.26* |
| **NPV** | 0.93±0.04 | 0.91±0.07† | 0.90±0.07* | 0.88±0.07† | 0.87±0.09† |
| **REM** | | | | | |
| **Accuracy** | 0.85±0.06 | 0.78±0.09† | 0.82±0.05* | 0.83±0.08* | 0.86±0.06 |
| **Sensitivity** | 0.74±0.16 | 0.51±0.24† | 0.52±0.16† | 0.63±0.23* | 0.63±0.16* |
| **Specificity** | 0.88±0.08 | 0.89±0.09 | 0.93±0.04 | 0.91±0.05 | 0.94±0.03 |
| **PPV** | 0.73±0.10 | 0.65±0.16* | 0.74±0.16 | 0.67±0.21 | 0.75±0.19 |
| **NPV** | 0.91±0.06 | 0.83±0.09† | 0.84±0.06† | 0.87±0.09* | 0.88±0.07* |
| **All Stages** | | | | | |
| **Sensitivity** | 0.73±0.06 | 0.57±0.09† | 0.62±0.08† | 0.63±0.11† | 0.64±0.03† |
| **Kappa** | 0.60±0.10 | 0.39±0.12† | 0.45±0.12† | 0.47±0.14† | 0.48±0.16† |

**indicates significantly different from Personalized p-value <0.05*
*†indicates significantly different from Personalized p-value <0.001*
